# The Prevalence and Factors Associated with Utilisation of Malaria Preventive Measures among Long Distance Travellers at Maunyamo Harbour in Mongu, Western Province of Zambia. A Cross-Sectional Study

**DOI:** 10.1101/2024.07.05.24310006

**Authors:** Inambao Chingumbe, Larry Lubinda Mooka, Mukumbuta Nawa

**Affiliations:** Department of Biostatistics and Epidemiology, School of Public Health and Environmental Studies, Levy Mwanawasa Medical University, P. O. Box 33991, Lusaka, Zambia

**Keywords:** Malaria, Preventive Measures, Long-Distance Travellers, Mongu Zambia

## Abstract

**Introduction:** Malaria is a significant public health concern in Zambia. Travelling is associated with exposure to different strains of malaria parasites whilst the use of preventive measures is not well documented among Zambia travellers. The Barotse flood plains in the upper Zambezi region with its swampy areas play a significant role in malaria transmission in the Western Province of Zambia. This study assessed the prevalence and factors associated with the utilisation of malaria preventive measures among long-distance travellers at Maunyamo harbour in Mongu district of Western Province.

**Methods:** This was a cross-sectional study design which targeted long-distance travellers from the Barotse flood plains who come to Mongu the capital and central business hub of Western Province. A sample size of 171 participants was selected using simple random sampling. Data was collected through a structured questionnaire with closed questions. Data was summarised using descriptive statistics such as frequency, percentages, and cross-tabulations with comparisons using Chi-Square or Fisher’s Exact test. Further, associations between the outcome variable and dependent variables were tested using univariate and multivariable logistic regression. A P-value of 0.05 was significant.

**Results:** A total of 171 respondents were included in the study, males 50.9% (87/171) and females 49.1% (84/171) were equally represented. A majority 74% (126/171) of participants were aware of malaria preventive measures, however, few actually used the conventional preventive measures such as mosquito repellents (43%), ITNs (19%), IRS (12%), and mosquito coils (5%) while a sizable number used unconventional means such as cutting and burning shrubs (18%). Factors associated with the use of conventional preventive measures included awareness of mosquito repellents as a preventive measure aOR 1.97 (P value < 0.001), whilst the younger age group of 21 – 25 years aOR 0.21 (P value 0.022) were significantly associated with less use of preventive measures compared to those aged 26 years and older. Factors such as sex, education levels and duration in business were not statistically significant.

**Conclusion:** Awareness of malaria preventive measures among long-distance travellers was relatively high but utilisation was much lower indicating a mismatch between knowledge and utilisation. Awareness of specific preventive measures such as mosquito repellents was associated with a higher utilisation rate whilst the younger age group was associated with lower utilisation. Socioeconomic factors like sex, education and duration were not statistically significant.

## Introduction

Malaria, a disease caused by an obligate intracellular protozoan parasite of the genus Plasmodium, is a serious mosquito-borne infectious disease, especially in sub-Saharan Africa, South America and Asia (1). In 2022, there were an estimated 249 million cases and 409,000 deaths worldwide and The African region bore 94% of the global malaria burden (2). In Zambia, there was an increase in reported malaria cases from 8.1 million cases in 2022 to 11.1 million cases in 2023 while deaths increased from 1343 in 2022 to 1602 in 2023 (3). The estimated incidence of malaria in Zambia was 340 cases per 1,000 populations per year while children under - five had a 29% prevalence based on rapid diagnostic tests (4). Malaria transmission occurs all year round, peaking in the late rainy season from February to May each year (5). Rural areas where 77% of Zambians reside face a 4.5 times greater risk than urban areas; the wetter, rural, and impoverished provinces of Luapula, Northern, Muchinga, North Western, and Western Provinces, along with adjacent rural areas of the Copperbelt and Eastern provinces, have the highest risk while Lusaka and Southern Province have the lowest risk (6).

The Barotse floodplains of the upper Zambezi River play a crucial role in malaria transmission in Western Province because the swampy areas of the flood plains serve as breeding habitats for mosquito vectors (7). Equally, the higher temperatures experienced in the province due to its lower altitudes also contribute to the rapid development and maturation of mosquito vector aquatic stages leading to higher vector populations leading to more exposure and infections (7). Malaria vector mosquitos in Zambia are known to exhibit sympathy with many species co-existing and transmitting malaria at the same time within the same locations including primary vectors such as *Anopheles funestus, an. gambiae* and *an. arabiensis* (8). People who reside in the flood plains are mainly occupied in fishing and agriculture, however, they rely on groceries and trade from Mongu which is the capital and commercial hub of Western Province (9). People who reside in the flood plains are therefore more exposed to malaria when they travel to the capital of Western Province Mongu for business, they also risk bringing different strains of the malaria parasites to the larger population of Mongu (10). Equally, the travellers risk exposure to other strains of the malaria parasites from other parts of the province and the country, so preventive measures against malaria among travellers is an important area that can help in the fight against malaria leading to the targeted elimination of malaria in Zambia by 2030 (11).

There is currently limited research on knowledge and practices regarding malaria among long-distance travellers from the Barotse flood plains in the Western Province of Zambia. Understanding the knowledge, and practices of long-distance travellers regarding malaria prevention is crucial for developing effective policies to protect the masses against malaria. This study therefore aimed at assessing the knowledge and practices of malaria prevention among long-distance travellers from the Barotse flood plains to Mongu the capital of Western province.

## Methodology

### Research Design

The study utilized a cross-sectional design.

### Study Setting

This study was done at Maunyamo harbour within Mongu district; the harbour is at the western edge of the business district of Mongu and it is where travellers from the Barotse flood plans stay when they come to do business in Mongu (12). The major activities which take place at this location include selling fish, sour milk, and reed mats which are produced in the flood plains (12). In return, they order groceries and other finished goods which they sell in the villages along the Zambezi River and its tributaries in the flood plains. The word *Maunyamo* is derived from the local language in Mongu called Lozi which means “a place of temporal residency”. Mongu is not only the capital of the Western province but also the commercial capital, so people from Kalabo, Sikongo, Nalolo and Senanga districts come to do business in Mongu. It was therefore selected to capture this demographic of people who constantly visit Mongu for business activities and go back to their villages in the flood plains along the Zambezi River and its tributaries.

### Population of the Study

The study focused on long-distance travellers who temporarily reside at Maunyamo Harbour in the Mongu district of Zambia’s Western province.

### Inclusion Criteria

The study included long-distance travellers who come to Mongu for business and go back to their villages in the flood plains. It only included those who were registered as temporal residents by the harbour authorities.

### Exclusion Criteria

Local residents who go to the harbour to sell merchandise were not included. Further, those who were too sick to answer questions were also excluded.

#### 3.1. Sample Size

The sample size was determined by the formula:

Sample Size n = N/1+Ne^2^

Where n = sample size, N = total population of 297 from the harbour authorities register, e = margin of error = 0.05 at 95% confidence level.

n = 171

#### 3.2. Sampling Techniques

A simple random sampling technique was used. From a register of 297 residents from the harbour master which was used as a sampling frame, Microsoft Excel was used to select the required number of 171.

### Data Collection

Data was collected using structured questionnaires that were administered by trained research assistants. The local language of Lozi was used to administer the questionnaires.

### Data Analysis

The questionnaires were coded and entered into Microsoft Excel. The data was cleaned to ensure accuracy and consistency. Frequencies of responses for categorical data and means for continuous data were calculated. Cross tabulations and Chi-Square were also calculated. Univariate and multivariable logistic regression was done to establish associations between the dependent and independent predictors.

### Ethical Considerations

Ethical clearance was obtained from Lusaka Apex Medical University Biomedical Ethics Committee IRB No. 00001131, FWA No. 00029892 and Protocol Approval No. 00734-24. Informed consent was obtained from all participants, and confidentiality, autonomy and respect were observed at all times.

## Results

The study included a total of 171 participants out of which 50.9% (87/171) were male and 49.1% (84/171) were females. The majority of participants were over 26 years old 62.7% (107/171), followed by those aged 21-25 years 20.5% (35/171), 16-20 years 16.4% (28/171), and only one was less than 15 years 0.6% (1/171). Participants’ education levels varied, with the highest proportion having attended University/College 50.3% (86/171), followed by those with Primary education 28.7% (49/171), Secondary education 18.7% (32/171), and those with no education were few 2.3% (4/171). Further, most participants had three or more years of experience in long-distance trade 69.6% (119/171), followed by those with at least two years of experience in long-distance trading 28.1% (48/171), and those with only one year’s experience in long-distance trading were few 2.3% (4/171). Moreover, the majority of the participants were aware of malaria interventions 73.7% (126/171), while a minority were not 26.3% (45/171). The most commonly used malaria interventions among participants were repellents 43.3% (74/171), followed by those who cut and smoked shrubs in their temporal homesteads 17.5% (30/171), while those who used ITNs were 19.3% (33/171), those who resided in temporal homestead which were sprayed with Indoor Residual chemicals were 12.3% (21/171), those who used mosquito coils were 4.7% (8/171) and least were those who used Intermittent Prophylactic Treatment 2.9% (5/171). Table 1 summarises the demographic characteristics and use of malaria prevention interventions during long-distance trading travels:

**Table 1:** Socio-demographic Profile of Respondents and Malaria Interventions Used.

| **Variables** | **Frequency (N=171)** | **Percentage (%)** |
| --- | --- | --- |
| **Sex** |  |  |
| Male | 87 | 50.9 |
| Female | 84 | 49.1 |
| **Age (years)** |  |  |
| < 15 | 1 | 0.6 |
| 16-20 | 28 | 16.4 |
| 21-25 | 35 | 20.5 |
| >26 | 107 | 62.6 |
| **Education level** |  |  |
| None | 4 | 2.3 |
| Primary | 49 | 28.7 |
| Secondary | 32 | 18.7 |
| University/College | 86 | 50.3 |
| **Experience in long-distance trade** |  |  |
| 1 year | 4 | 2.3 |
| 2 years | 48 | 28.1 |
| 3 years and above | 119 | 69.6 |
| **Malaria intervention awareness** |  |  |
| Yes | 126 | 73.9 |
| No | 45 | 26.3 |
| **Malaria interventions** |  |  |
| Repellents | 74 | 43.3 |
| ITNs | 33 | 19.3 |
| Mosquito coils | 8 | 4.9 |
| IPT | 5 | 2.9 |
| IRS | 21 | 12.3 |
| Cutting shrubs | 30 | 17.5 |
| **Access to malaria interventions** |  |  |
| Yes | 125 | 73.1 |
| No | 46 | 26.9 |

| **Comfortable using the malaria interventions** |  |  |
| --- | --- | --- |
| Yes | 0 | 0 |
| No | 171 | 100 |

### Common Challenges in Protecting Against Malaria

The most common challenges faced by participants in protecting themselves against malaria included the lack of availability of preventive measures 45 (26.3%), the cost associated with these preventive interventions 40 (23.4%), a lack of knowledge about malaria preventive measures 25 (14.6%), and the inconvenience of carrying preventive measures while travelling 15 (8.8%).

### Factors Associated with the Use of Malaria Preventive Measures while Away from Home

The study showed that the younger age group 20 – 25 years old were less likely to use malaria preventive measures aOR 0.21 (P-value = 0.022) compared to older people in the age group 26 years and above. On the other hand, those who were aware of mosquito repellants were more likely to use malaria preventive measures aOR 1.97 (P-value < 0.001) compared to those who knew about other measures such as IRS and ITNs during long-distance travels. However, there were no statistically significant differences between males and females, by educational levels attained and years of experience in long-distance trading. Table 3 summarises the factors associated with the use of malaria preventive measures among long-distance traders from the Barotse flood plains in Mongu, Western Province:

**Table 2:** Common challenges mentioned in protecting against malaria.

| **Challenges** | **No.** | **Percentage%** |
| --- | --- | --- |
| Lack of availability | 45 | 26.3 |
| Cost | 40 | 23.4 |
| Lack of knowledge | 25 | 14.6 |
| Inconvenience in carrying measures | 15 | 8.8 |

**Table 3:** Factors Associated with the Use of Malaria Preventive Measures.

| Variable | Category | uOR | P Value | aOR | P Value |
| --- | --- | --- | --- | --- | --- |
| Age | < 15 yrs. | 1 |  | 1 |  |
|  | 16-20 yrs. | 1.12 | 0.823 | 0.28 | 0.073 |
|  | 21 – 25 yrs. | 1.07 | 0.872 | **0.21** | **0.022\*** |
|  | 26 yrs & above | Ref |  | Ref |  |
| Sex | Male | Ref |  | Ref |  |
|  | Female | 3.67 | < 0.001 | 1.11 | 0.859 |
| Educational level | None | Ref |  | Ref |  |
|  | Educated | 3.82 | < 0.001 | 1.79 | 0.407 |
| Awareness of Malaria preventive measures | Repellants | **2.0** | **< 0.001\*** | **1.97** | **< 0.001\*** |
|  | Others (IRS, ITN) | Ref |  | Ref |  |
| Experience in long-distance trade | 1 year | Ref |  | Ref |  |
|  | 2 years | 0.15 | 0.115 | 0.18 | 0.258 |
|  | 3 years and more | 3.27 | 0.322 | 1.15 | 0.949 |

\*Statistically significant variables

## Discussion

This study set out to find the knowledge and use of malaria preventive measures among long-distance travellers from the Barotse flood plains at Maunyamo harbour in Mongu district and factors associated with the use of malaria preventive measures. The study found that there were high levels of awareness of malaria preventive measures among long-distance traders in Mongu district of Western province, however, the use of malaria preventive measures was low as less than half of the participants used at least any preventive measures such as repellents, ITNs, IRS or prophylaxis. The findings of this study are similar to the findings of a recently published study in Mansa district of Luapula province in Zambia which found that out of 96 people with high knowledge of malaria prevention, only 10.4% (10/96) practised the preventive measures (13). Similarly, another study of long-distance travellers also found high levels of knowledge of malaria but low utilisation of preventive measures (14). The use of interventions seems to be generally low even in other areas such as COVID-19 vaccinations (15). The discrepancy between knowledge and practice may be multifactorial; in our study, young persons aged 21 to 25 years were associated with lower utilisation of preventive practices compared to older persons above 26 years. In high transmission settings such as Zambia, the risk perception of malaria among adults is low because they have partial immunity to symptomatic malaria or adults may only experience mild symptoms when infected compared to under-five children (16). Because they seldom fall ill due to malaria even when they are bitten by mosquitoes, their risk perception is low and are therefore not motivated to use malaria preventive measures therefore creating a psychological barrier to practising malaria prevention (17). Even when infected, they usually have subclinical parasitaemia and so when bitten by mosquito vectors, they will pass the infections to others including under-five children and pregnant women who are prone to symptomatic and severe malaria leading to adverse outcomes and death (17). Therefore, sensitisation and messaging of malaria prevention for young adults and older people must be not only for personal prevention but also to cut transmission cycles to prevent malaria in the most vulnerable groups such as under-five children and pregnant women. Other than the low perception of malaria risk, other factors included lack of availability of preventive measures, cost associated, lack of knowledge and inconvenience of carrying products such as ITNs.

Interventions such as Mass Distribution of ITNs and yearly spraying of residual chemicals usually target universal coverage of households and do not cover those who are travelling, so a man may leave his spouse and children using ITNs at home that the household was given during mass distribution campaigns and be exposed to mosquito bites when he travels out of town for business (18). The findings of this study are similar to what was reported in a systematic review of 30 studies done in Low and Middle-Income Countries (LMIC) which categorised barriers to malaria prevention into lack of geographical accessibility, affordability, availability and acceptability (18). Therefore, interventions targeting domestic travellers within high transmission settings such as Zambia are required as current interventions such as chemoprophylaxis only target international travellers from non-malaria endemic areas travelling to malaria-endemic areas (19). In fact, chemoprophylaxis against malaria among domestic travellers within malaria-endemic areas is discouraged as it can lead to the development of resistance to commonly used drugs (20). In high transmission settings such as Zambia, the use of mosquito repellents, coils and ITNs are good options for domestic travellers to protect themselves against mosquito bites when they travel, however, they are not part of the interventions given to people on a large scale specifically targeting domestic travellers.

Further, this study found that knowledge of some malaria preventive measures particularly mosquito repellents was associated with increased usage of malaria preventive measures among long-distance travellers. This is different from what was found in another study in Cambodia which found that there was high knowledge and acceptability of mosquito repellents, however, their use was low (21). Our study therefore can be used to inform policies on increasing the use of mosquito repellents in the sub-Saharan African settings among vulnerable populations such as long-distance travellers and refugees. A recent Cochrane systematic review and meta-analysis found that mosquito repellents were effective in reducing falciparum malaria incidence IRR 0.74 (95%CI 0.56 – 0.98) and prevalence OR 0.81 (0.67 – 0.97) (22). The review further indicated that repellents can make a difference in protecting especially high-risk groups where other conventional interventions such as ITNs and IRS cannot be deployed such as travellers, refugees and forest goers (22). Topical mosquito repellents emit volatile substances that irritate the olfactory systems and trigger avoidance behaviour among mosquito vectors (23). Further, the use of mosquito repellents among malaria-at-risk groups who may not benefit from ITNs and IRS such as refugees and travellers as they are mobile is also associated with the prevention of other vector-borne diseases such as Yellow fever, Dengue fever and Chikungunya among others (23).

In addition, our study did not find significant statistical differences in the use of malaria preventive measures by sex among long-distance travellers. Other studies among travellers also did not find significant differences between male and female travellers (24, 25). This indicates no significant sex inequities in the use of malaria preventive measures among long-distance travellers. Further, educational levels were also not statistically significant in our study, but another study found that higher education was associated with noncompliance with malaria preventive measures (24). It may be expected that higher education would be associated with more use of malaria preventive measures during long-distance travels, however, our study showed that it was not significant probably because education level in general is a distal variable to knowledge while we asked a more specific proximal question on knowing specific malaria preventive measures which was factored in the multivariable model and reduced the confounding rendering education not to be significant. There is evidence of confounding because the education level variable was initially significant in the univariate logistic regression model but became insignificant when we adjusted for other variables including knowledge of specific preventive measures; in the multivariable model, when we removed knowledge of specific measures, education level became significant again. Further, the number of years in long-distance trade was also not statistically significant in our study. We did not find literature on the association between the number of years in long-distance trade and the risk of malaria. It may be expected that those with many years of trading would use more preventive measures compared to those with fewer years in long-distance trading; however, this was not the case in our study and more studies are needed to understand why the longer duration of long-distance trade may not be associated with use o of malaria preventive measures.

### Study Limitations

The study was done in one place, a harbour in Mongu that hosts people who travel from the flood plains in Mongu, so it is not generalizable to other domestic travellers within Zambia or international travellers.

## Conclusion

This study found that the prevalence of the use of malaria preventive measures among long-distance travellers in Mongu was low, however, awareness was high. Factors associated with low utilisation include younger age groups while knowledge of preventive measures increased the likelihood of use of preventive measures. Factors such as sex and education levels were not significantly associated with the use of prevention measures for malaria.

### Recommendations

The study recommends sensitisation of long-distance travellers in malaria prevention measures to create more awareness of malaria prevention as this was positively associated with the use of preventive measures. Further, the study recommends targeting young adults who have a low-risk perception of malaria because of partial immunity that the risk is not just for them but the community because mosquitos that bite them can also bite younger children and pregnant women who can develop severe disease, so they need to have a sense of community to prevent malaria even if they may not fall sick themselves. We further recommend sensitisation and roll out of traveller-friendly malaria preventive measures such as repellents and mosquito coils for domestic travellers.

## Data Availability

All data produced in the present study are available upon reasonable request to the authors

